# Perfusionist nursing as a key element in organ preservation and viability in uncontrolled DCD (uDCD) after failed ECPR: experience and outcomes of transplanted organs

**DOI:** 10.64898/2026.02.16.26346412

**Authors:** Mireia Gispert Martínez, Marina Chordà Sánchez, Oriol Roselló Castells, Ángel Ruiz Arranz, Jordi Castillo Garcia

**Affiliations:** Hospital Clínic de Barcelona; Diputació de Barcelona; Hospital de Bellvitge

**Keywords:** Uncontrolled Donation after Circulatory Death, Maastricht II, Cardiorespiratory Arrest, Normothermic Regional Perfusion, ECMO, perfusion, organ donation, transplantation

## Abstract

**Objective:** To analyze the experience of the last six years with ECMO in Uncontrolled Donation after Circulatory Death (uDCD), assessing the clinical and logistical factors that determine donation effectiveness and the viability of retrieved organs, with the nurse perfusionist as the central figure in organ perfusion.

**Methods:** Retrospective observational study of uDCD procedures performed at Hospital Clínic de Barcelona between June 2019 and October 2025.

**Results:** Of 184 out-of-hospital ECMO-CPR activations, 108 (58.7%) underwent perfusion; 72 donor cases (66.7%) were generated, and 109 kidneys (75.7%) and 3 livers (4.15%) were retrieved. The annual number of uDCD donors was heterogeneous. Compared with non-effective donors, effective donors were significantly younger (48.1 ± 12.4 vs 53.0 ± 10.7 years, p=0.03) and had fewer comorbidities such as hypertension (13.8% vs 33.0%, p=0.018) and diabetes (4.1% vs 16.6%, p=0.027). Although effective donors had a shorter cannulation time (25.6 ± 13.9 vs 29.1 ± 11.9 min, p=0.09), the difference was not statistically significant; however, cardiocompressor time did show a significant difference (58.9 ± 17.7 vs 65.8 ± 18.2 min, p=0.03).

**Conclusions:** uDCD was a useful source of transplantable organs, mainly kidneys (two out of every three perfused patients became donors), in the current context of scarcity of brain-dead donors. Shorter warm ischemia times (cardiocompressor and cannulation times) were significantly associated with more effective organ donation. The multidisciplinary transplant team may benefit from perfusion professionals with expertise in extracorporeal oxygenation therapy.

## Introduction

In recent years, a progressive decline in organ donors with brain death has been observed^1^. In this context, the search for alternative sources of organs has become an important task for transplant organizations. In Spain, the first published cases of Maastricht type III donors (controlled donation after circulatory death following the limitation of life-sustaining treatment) appeared in 2011^2^, and they have now surpassed donors with brain death^1^.

However, it remains necessary to continue making progress to avoid donor shortages, and work is underway with patients suffering from both in-hospital and out-of-hospital cardiac arrest (CA) in whom cardiopulmonary resuscitation (CPR) maneuvers and the implantation of an extracorporeal membrane oxygenation (ECMO) system prove unsuccessful: uncontrolled donation after circulatory death (uDCD), Maastricht type II^3^, which has become an effective alternative to increase the availability of organs for transplantation^1^.

The uDCD process involves several healthcare organizations, including Emergency Medical Systems (both field teams and coordination centers), as well as the hospital team available 24 hours a day. In these cases, warm ischemia times are prolonged, and up to 150 minutes are permitted before the initiation of ECMO. In such donors the heart is not viable, while the lungs may be suitable if the receiving center has a lung transplant program.

The first modern uDCD protocols were developed in the late 1980s, almost simultaneously, at Hospital Clínico San Carlos (Madrid), the A Coruña University Hospital Complex (CHUAC), and Hospital Clínic de Barcelona^4^ (HCB). HCB implemented its in-hospital uDCD program (Maastricht IIa) in 1985 and, since 2003, has managed out-of-hospital activations involving ECMO-CPR (Maastricht IIb). However, organ perfusion with ECMO at HCB was not carried out by experts in Extracorporeal Circulation Techniques (“perfusion nurses”) until 2019, which increased the safety of the procedures.

The aim of this study was to analyze the experience with Maastricht type II uDCD over the last six years at this university hospital, examining the clinical and logistical factors that influence donation effectiveness and the viability of retrieved organs, highlighting the role of the perfusion nurse as the central figure in organ perfusion.

## Methods

A retrospective observational study was conducted on uncontrolled donation after circulatory death (uDCD) cases performed at Hospital Clínic de Barcelona between June 2019 and October 2025. The study was approved by the hospital’s Ethics Committee (Record: HCB/2023/1124).

Patients aged 14 to 64 years who suffered an out-of-hospital cardiac arrest (OHCA) and were initially activated under the ECMO-CPR protocol were included, provided that advanced cardiopulmonary resuscitation (CPR) maneuvers were unsuccessful and they were transported to the hospital for death certification. Patients were excluded if they presented with hypothermia (temperature below 32 °C), injuries incompatible with donation, judicial or family refusal, or if they ultimately entered the ECMO-CPR protocol. Patients exceeding the temporal limits established in the protocol were also excluded, including those with a no-flow time (absence of advanced life support) greater than 15 minutes and/or a total warm ischemia time—defined as the interval between cardiac arrest and initiation of post-mortem ECMO—exceeding 150 minutes.

The main variables included sociodemographic data, medical history and comorbidities, activations initiated by the transplant coordination team, perfusion activation, transplanted organs, chest compression duration, and cannulation times. Data were collected using an ad hoc questionnaire completed through a review of medical records.

Descriptive statistical analysis was expressed as mean and standard deviation for quantitative variables and as frequencies and percentages for qualitative variables in cases of normal distribution. Statistical comparisons between groups were performed using Student’s t-test for quantitative variables and the chi-square test for qualitative variables. A p-value of ≤ 0.05 was considered statistically significant. Statistical analysis was performed using SPSS for Windows, version 21.

## Results

During the 2019–2025 period, 184 out-of-hospital ECMO-CPR activations were recorded at Hospital Clínic de Barcelona, of which 108 patients received ECMO therapy upon arrival to the hospital for Maastricht IIb uDCD. Of the 184 activations, 108 (58.7%) underwent perfusion, generating 72 donor cases (66.7%) and yielding 109 kidneys (75.7%) and 3 livers (4.15%).

The annual number of uDCD donors varied throughout the study period, with approximately nine male donors for every female donor. The overall mean donor age was 50.5 ± 10.3 years, and donors of both sexes presented with grade II overweight: BMI 26.9 ± 5.3 in women and 28.4 ± 4.2 in men (Table 1).

**Table I.** Distribution and characteristics of donors over the 7-year study period.

| Variable | 2019 | 2020 | 2021 | 2022 | 2023 | 2024 | 2025 | Total |
| --- | --- | --- | --- | --- | --- | --- | --- | --- |
| Activated cases | 52 | 10 | 13 | 45 | 31 | 22 | 11 | 184 |
| Perfusion activation | 21<br>(40.4%) | 6 (60%) | 7<br>(53.8%) | 28<br>(62.2%) | 24<br>(77.4%) | 14<br>(63.6%) | 8<br>(72.7%) | 108<br>(58.7%) |
| Non-effective donors | 1<br>(4.8%) | 1<br>(16.7%) | 2<br>(28.6%) | 12<br>(42.9%) | 14<br>(58.3%) | 6<br>(42.9%) | 0 (0%) | 36<br>(33.3%) |
| Effective donors | 20<br>(95.2%) | 5<br>(83.3%) | 5<br>(71.4%) | 16<br>(57.1%) | 10<br>(41.7%) | 8<br>(57.1%) | 8<br>(100%) | 72<br>(66.7%) |
| Kidneys transplanted | 20<br>(50%) | 7 (70%) | 6 (60%) | 32<br>(100%) | 20<br>(100%) | 10<br>(62.5%) | 14<br>(87.5%) | 109<br>(75.7%) |
| Livers transplanted | 0 (0%) | 0 (0%) | 1 (20%) | 1<br>(6.25%) | 1 (10%) | 0 (0%) | 0 (0%) | 3<br>(4.15%) |
| Age (mean ± SD) | 50.6<br>(12.7) | 57.7<br>(5.7) | 42.9<br>(11.8) | 48.1<br>(9.8) | 51.2<br>(7.7) | 48.7<br>(12.3) | 53.0<br>(8.0) | 50.1<br>(10.3) |
| Male sex, n (%) | 18<br>(90%) | 4 (80%) | 3 (60%) | 14<br>(87.5%) | 9 (90%) | 5 (83%) | 8<br>(100%) | 60<br>(83.3%) |
| Male BMI (mean ± SD) | 27.5<br>(3.4) | 31.0<br>(6.6) | 27.0<br>(2.6) | 29.1<br>(4.6) | 28.6<br>(4.1) | 27.4<br>(2.8) | 27.6<br>(5.0) | 28.4<br>(4.2) |
| Female BMI (mean ± SD) | 29.7<br>(7.7) | 25.7 | 30.2<br>(4.1) | 27.9<br>(6.5) | 26.3<br>(2.1) | 20.4<br>(5.4) | — | 26.9<br>(5.3) |

Abbreviation: BMI = body mass index.

The main causes of death among perfused donors (perfusion activation) were ischemic heart disease in 68 cases (63%), cardiac arrhythmias in 23 cases (21.3%), and other causes in 17 cases (15.7%), including pulmonary thromboembolism, aneurysms, aortic dissections, and others.

Compared with non-effective donors, effective donors were statistically significantly younger (48.1 ± 12.4 vs. 53.0 ± 10.7 years, p = 0.03) and had fewer comorbidities such as hypertension (13.8% vs. 33%, p = 0.018) and diabetes (4.1% vs. 16.6%, p = 0.027). Although effective donors had shorter cannulation times (25.6 ± 13.9 vs. 29.1 ± 11.9 minutes, p = 0.09), the difference was not statistically significant.

However, there were significant differences in the duration of mechanical chest compressions (58.9 ± 17.7 vs. 65.8 ± 18.2 minutes, p = 0.03). Table 2 summarizes the variables associated with effective donation.

**Table II.**
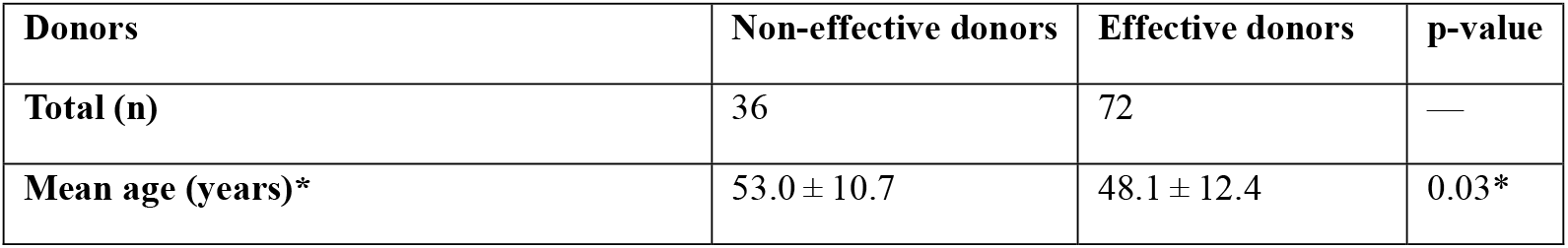
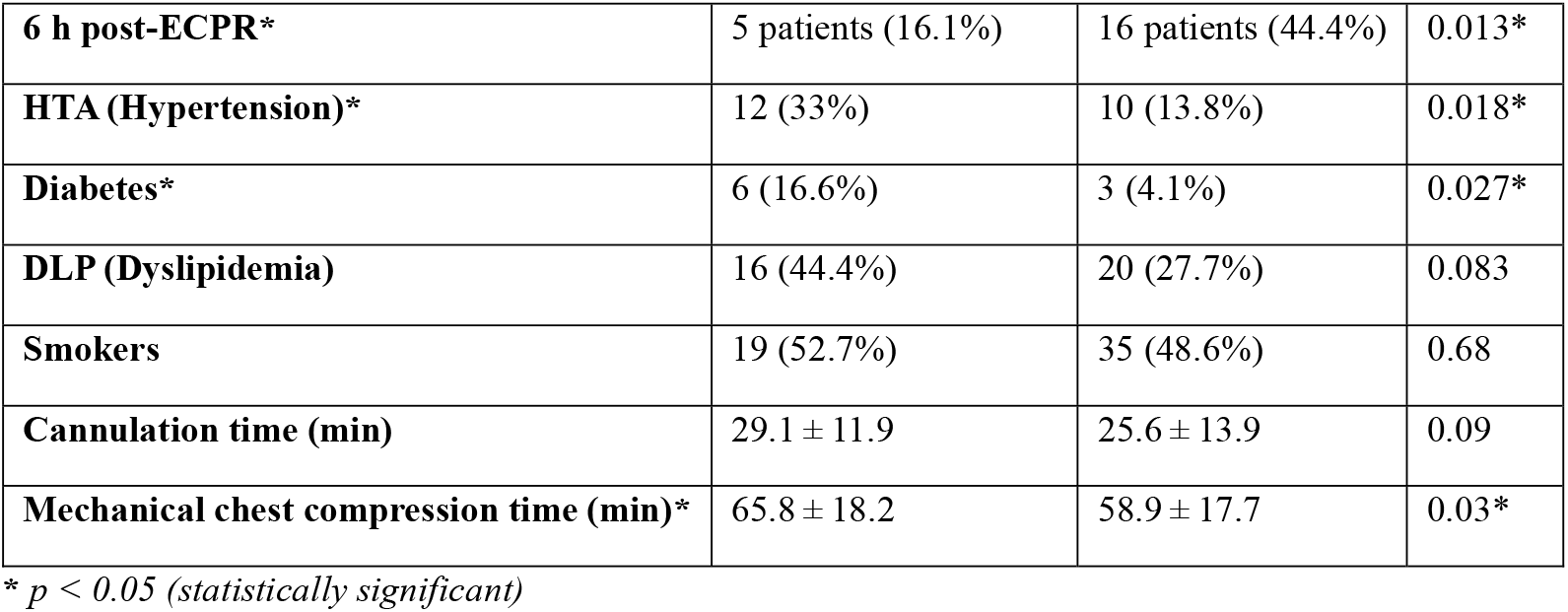
Relationship between study variables and effective organ donation.

| Donors | Non-effective donors | Effective donors | p-value |
| --- | --- | --- | --- |
| Total (n) | 36 | 72 | — |
| Mean age (years)* | $53.0 \pm 10.7$ | $48.1 \pm 12.4$ | 0.03* |
| **6 h post-ECPR\*** | 5 patients (16.1%) | 16 patients (44.4%) | 0.013* |
| **HTA (Hypertension)\*** | 12 (33%) | 10 (13.8%) | 0.018* |
| **Diabetes\*** | 6 (16.6%) | 3 (4.1%) | 0.027* |
| **DLP (Dyslipidemia)** | 16 (44.4%) | 20 (27.7%) | 0.083 |
| **Smokers** | 19 (52.7%) | 35 (48.6%) | 0.68 |
| **Cannulation time (min)** | 29.1 ± 11.9 | 25.6 ± 13.9 | 0.09 |
| **Mechanical chest compression time (min)\*** | 65.8 ± 18.2 | 58.9 ± 17.7 | 0.03* |

\* $p < 0.05$ (statistically significant)

## Discussion

The results obtained during the 2019–2025 period confirm that the conversion of a failed ECMO-CPR protocol into uDCD is feasible and has enabled transplantation of 75.7% of the kidneys recovered. Our study aligns with current literature regarding favorable prognostic factors, such as the finding that effective donors had a mean age of 48 years. Although recent publications suggest that donors should ideally be younger than 60 years^5^, stricter international guidelines prioritize donors under 45 years of age and consider those older than 45 with greater caution^6^.

Consistent with the existing literature, our optimal donor profile is a young adult, normotensive, non-diabetic, and without cardiovascular disease^7^. We also concur that hypertension, diabetes, dyslipidemia, and smoking are markers of increased coronary artery disease risk, which negatively affects graft survival^6^. Similarly, other Spanish uDCD series^8^ and European reviews^9^ describe donation after circulatory death as a highly time-dependent process in which rapid initiation and limitation of chest compressions, as well as timely cannulation to begin preservation, are essential to minimizing warm ischemia and reducing the incidence of early and primary graft dysfunction—one of the limitations of our study.

uDCD is a procedure of high technical and logistical complexity, requiring immediate multidisciplinary coordination and continuous availability, which partly explains the variability in the number of cases across the study years. Moreover, it demands particularly complex hemodynamic and transfusion management during regional perfusion, often in the setting of post-mortem vasoplegia, with the aim of maintaining adequate mean arterial pressure, sufficient hematocrit, and stable acid–base balance to ensure effective perfusion of abdominal organs, especially the liver and kidneys.

Recent reviews and international guidelines emphasize that normothermic regional perfusion should only be performed in centers equipped with teams specifically trained in extracorporeal oxygenation and perfusion techniques, including perfusionists and ECMO specialists. They also recommend mobile team models to support hospitals without this expertise or equipment^3^,^10^.

One limitation of our study is the absence of data prior to the incorporation of perfusionists into the uDCD process. Consequently, it is not possible to compare outcomes before and after the introduction of this role—an analysis that would have been of particular interest. Future studies aim to specifically examine early and primary graft dysfunction in this therapeutic context, with the objective of providing additional evidence on its impact on post-transplant outcomes.

In conclusion, Maastricht type II uDCD represents a robust and ethically sound source of organs— primarily kidneys—in the current context of decreasing brain-death donation. More than half of activations (58%) resulted in uDCD cases, and shorter warm ischemia times (chest compression and cannulation times) were associated with more effective organ donation.

## Data Availability

De-identified clinical data supporting the findings of this study are not publicly available due to institutional and ethical restrictions, but are available from the corresponding author upon reasonable request and subject to approval by Hospital Clínic de Barcelona.

## Acknowledgments

We would like to thank the perfusionists of Hospital Clínic for their commitment and daily work.

